# Factors preventing SARS-CoV-2 transmission during unintentional exposure in a GP practice: a cohort study of patient contacts; Germany, 2020

**DOI:** 10.1101/2021.03.17.21251046

**Authors:** T. Sonia Boender, Jennifer K. Bender, Angelika Krüger, Kai Michaelis, Udo Buchholz

## Abstract

Two general practitioners (GPs) a with SARS-CoV-2 infection provided in-person patient care to patients of their joint medical practice before and after symptom onset, up until SARS-CoV-2 laboratory confirmation. In a retrospective cohort study of patient contacts, we assessed the risk (frequency and determinants) of SARS-CoV-2 transmission from the GPs to their patients. Our findings support the use of facemasks for GPs, and short consultation time, to minimize the risk of transmission.

**Summary:** Two general practitioners (GPs) with SARS-CoV-2 infection provided in-person patient care to patients of their joint medical practice before and after symptom onset, up until SARS-CoV-2 laboratory confirmation. Through active contact tracing, the local public health authorities recruited the cohort of patients that had contact with either GP in their putative infectious period. In this cohort of patient contacts, we assess the frequency and determinants of SARS-CoV-2-transmission from GPs to patients. We calculated incidence rate ratios (IRR) to explore the type of contact as explanatory variable for COVID-19 cases. Among the cohort of 83 patient contacts, we identified 22 (27%) COVID-19 cases including 17 (21%) possible, 3 (4%) probable, and 2 (2%) confirmed cases. All 22 cases had contact with a GP when the GP did not wear a mask, and/or when contact was ≥10 minutes. Importantly, patients who had contact <10 minutes with a GP wearing a facemask were at reduced risk (IRR 0.21; 95%CI 0.01-0.99) of COVID-19. This outbreak investigation adds to the body of evidence in supporting current guidelines on measures at preventing transmission of SARS-CoV-2 in an outpatient setting.

## Text

### Background

In Germany, the first case of COVID-19 was confirmed on 27 January 2020 (1). This was followed by several introductions and clusters of cases of COVID-19. On 27 February, Germany reported a total of 26 cases (2). By 3 March 2020, 196 cases were reported, of which the majority (n=101) belonged to a large cluster in North-Rhine Westphalia or were linked to importation from high risk areas (n=35, from Italy, China, Iran) (3). Three days later, on 6 March 2020, a total of 639 confirmed COVID-19 cases were reported in Germany (4). Extensive contact tracing by the responsible health authorities was ongoing.

In this field report, we report about one of the first of ten cases of COVID-19 and the associated cluster of contacts and cases that was notified to a local public health authority in Germany (district population of >500.000), early March 2020, in the context of a GP practice. In addition to contact tracing activities, we conducted an outbreak investigation to assess the likelihood of transmission in a GP setting by means of a retrospective cohort study of patient contacts.

### Epidemiology of the incident

Two GPs, working in their joint medical practice, had fallen ill with COVID-19 end of February / early March 2020. To ensure confidentiality, we report an anonymised timeline, counting days 1 to 22, starting two days before the first day of symptom onset of one of the GPs. Both GPs had been attending to patients shortly before and after onset of respiratory symptoms, until laboratory confirmation of SARS-CoV-2 infection on day nine (**Figure 1**). Of note, because of the limited number of cases of COVID-19 in the area at that time, exposure to SARS-CoV-2 outside of the GP practice was deemed unlikely.

**Figure 1.**
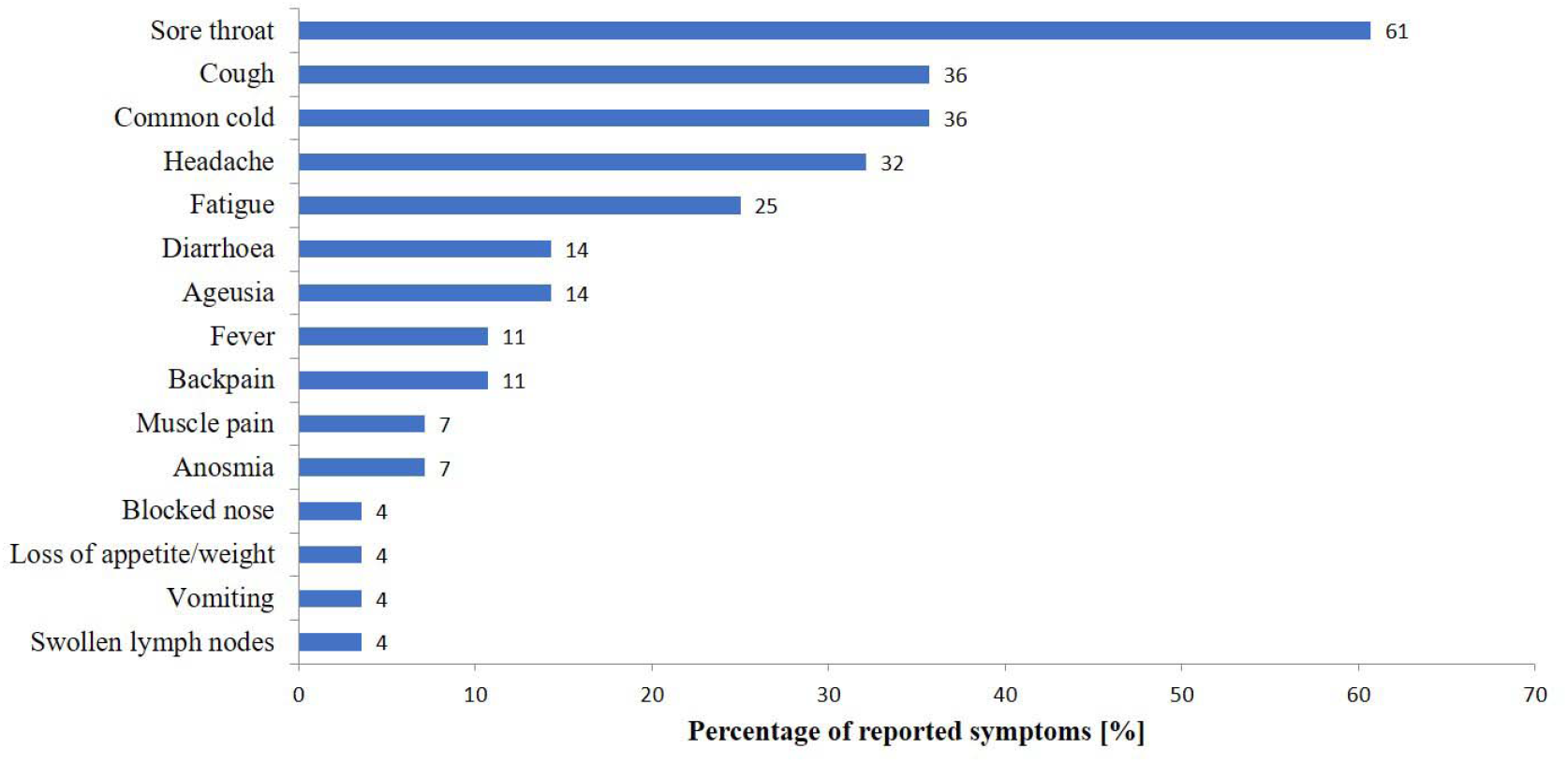
Percentage of self-reported onset of COVID-19-associated symptoms after contact with either GP, presented as a percentage of the full cohort, as recalled by the 28/83 study participants.

### Response & outbreak investigation

Through active contact tracing as by local guidelines, in line with national and international guidelines (5, 6), close contacts were identified, interviewed by phone, and ordered quarantine and PCR-testing, if applicable. For the purpose of the analytical study (outbreak investigation), the local public health authorities recruited the cohort of patients that had contact with either GP in their putative infectious period (starting two days before and ending 10 days after the date of symptom onset). The cohort of patients that was exposed to SARS-CoV-2 during their GP visit provided the unique opportunity to investigate the likelihood of transmission in a GP setting. Therefore, and in accordance with national contact tracing guidelines (5), we included patients in our cohort who were exposed to either GP two days before their respective date of symptom onset until laboratory confirmation of SARS-CoV-2 infection of the GP. Thus, the study period included all valid patient contacts from day one to day nine of the anonymised timeline. The GPs were self-isolating from day nine onwards. Both GPs made a full recovery. Close contacts of either GP were ordered a 14-day home quarantine. Quarantine included active daily monitoring of COVID-19 symptoms and testing for SARS-CoV-2 in the event of symptom onset and/or at the end of the quarantine period. The full referent period can therefore be defined as the day one (two days before the symptom onset of the first GP) up to and including day 22 (last day of quarantine). Work-related contacts included six practice assistants; all reported distant contact, tested negative for SARS-CoV-2 and were not considered part of the cohort.

In addition to routine contact tracing phone calls, we interviewed all patient contacts by phone at the end of their putative incubation period (14 days after exposure) and again four weeks later, using a standardised questionnaire. We collected information on demographics, medical and travel history. In addition, for each GP visit, the (medical) purpose, duration and type of contact with the GP was extracted, including which GP was seen, the distance to the GP, if the patient was physically examined, and whether or not the GP wore a medical facemask. At the time, wearing a facemask by GPs was not yet recommended as a standard infection prevention and control measure for all situations of direct contact to all patients (regardless of infection status) in Germany (7).

We collected information on potential symptom onset in the 14 days after risk contact with the GP, including the type and timing of general and COVID-19-associated symptoms (8, 9). New onset was defined as a symptom that started the day after the risk contact up until 14 days after the GP visit. COVID-19-compatible symptoms were separated into respiratory and non-respiratory symptoms / clinical diagnoses. Respiratory symptoms / clinical diagnoses included cough, sore throat, common-cold (runny nose), shortness of breath and/or pneumoniae. Non-respiratory symptoms included fever, anosmia (loss of the sense of smell), ageusia (loss of the sense of taste), dysgeusia (distortion of the sense of taste), headache, back-pain, muscle ache, joint-pain, loss of appetite and/or weight, nausea, vomiting, diarrhoea, conjunctivitis, skin rash, swollen lymph nodes and/or apathy.

Case definitions were based on self-reported symptoms and laboratory criteria. Possible cases reported onset of at least two COVID-19-compatible symptoms. Probable cases met the criteria for a possible case, but also had pneumonia, anosmia, ageusia or dysgeusia. Confirmed cases tested positive for SARS-COV-2, regardless of symptoms (i.e. national COVID-19 case definition).

We used negative binomial regression to calculate incidence rate ratios (IRR with 95% confidence intervals [95%CI]) to explore the risk (frequency and determinants) of SARS-CoV-2 transmission from GPs to patients using the MASS and fmsb packages in R Version 3.6.3.

We conducted this outbreak investigation as part of the official tasks of the local public health authorities of the respective district supported by the RKI upon official request in accordance to §4 of the German Protection against Infection Act. Therefore, this investigation was exempt from additional institutional review.

## Results & discussion

We reached 91 (70%) of 131 patient contacts as traced by the local health authority. The 40 patient contacts who could not be reached had a median age of 32 [IQR 28-43] years, and 21 (53%) were female. Of the 91 reached, two contacts did not meet our inclusion criteria --they were not exposed to or were exposed outside the infectious period of either GP--and six did not consent. Thus, 83 (91%) of 91 were included in the cohort study.

The median age was 45 [IQR 7-87] years, 56 (68%) were female. There were 25 (30%) current smokers (tobacco) and 54 (65%) reported underlying conditions. Fifteen people reported traveling in the two weeks before visiting their GP; no one travelled to areas classified as risk areas (international) or particularly affected areas (in Germany). Some patients visited their GPs more than once during the infectious period of the GPs, we therefore recorded 89 contact-events in our study (GP_X_ n=43; GP_Y_ n=46). Patients spent a median of 10 minutes (IQR 10-20) with their GP. The GPs wore a medical facemask during 31 (35%) of 89 contact-events, more often during physical examination (25/56; 45%) compared to no physical examination (6/32; 19%; Fisher’s exact *P*=.046).

In total, 28 participants (33.7%) reported onset of COVID-19 associated symptoms after exposure to either GP, 22 (26.5%) reported respiratory symptoms including fever, 21 (25.3%) reported respiratory symptoms, irrespective of fever. Sore throat (n=17; 61%), cough (n=10; 36%), common cold (n=10; 36%), headache (n=9; 32%) and apathy (n=7; 25%) were the most frequently reported symptoms (**Figure 1**). We identified 22 (27%) COVID-19 cases, including 17 (21%) possible, 3 (4%) probable, and 2 (2%) confirmed cases (**Figure 2 & Table 1**).

**Figure 2.**
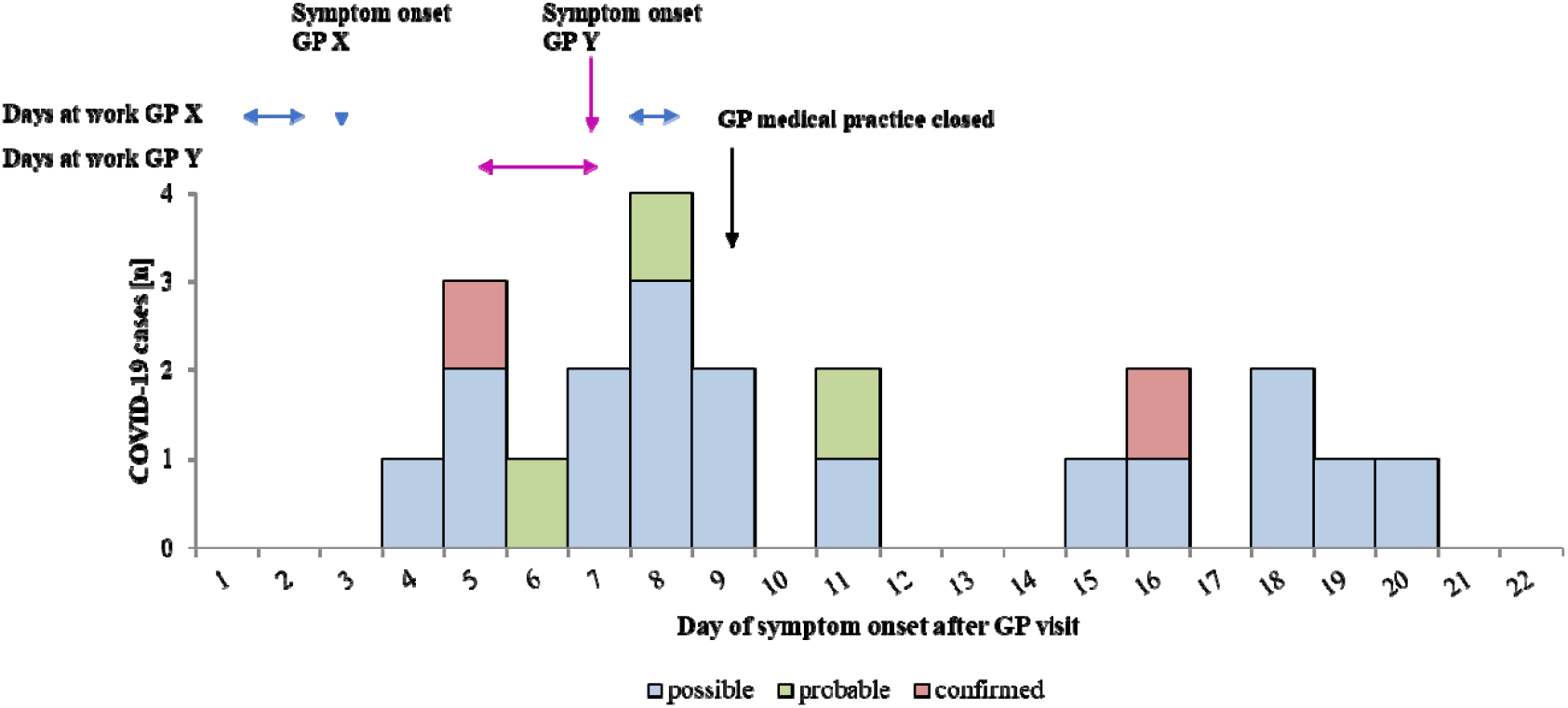
Date of symptom (following the anonymized timeline) onset of possible, probable and confirmed COVID-19 cases within the cohort of patient contacts of two SARS-CoV-2-positive GPs in a medical practice in Germany, February-March 2020. Symptom onset of either GP, the days they attended to patients, and closure of the GP medical practice are marked by blue (GP_X_), pink (GP_Y_) and black arrows, respectively.

**Table 1.** Summary of characteristics of the five confirmed and probable COVID-19 cases within the cohort of patients contacts of two SARS-CoV-2-positive GPs in a GP practice in Germany, 2020.

|  | Case 1 | Case 2 | Case 3 | Case 4 | Case 5 |
| --- | --- | --- | --- | --- | --- |
| Case classification | Confirmed | Confirmed | Probable | Probable | Probable |
| Age group (years) | Between 20-25 | Between 40-45 | Between 20-25 | Between 45-50 | Between 80-85 |
| Sex | Female | Female | Female | Female | Female |
| Incubation period (days) | 3 | 14 | 3 | 1 | 5 |
| Underlying conditions | No | No | No | Yes | Yes |
| GP visited | X | X | Y | Y | Y |
| Time with GP (minutes) | 10 | 10 | 13 | 12 | 5 |
| Distance to GP (meter) | <1 | <1 | 1-2 | <1 | <1 |
| Close contact<sup>a</sup> | Close | Close | Close | Close | Close |
| Physical examination by GP | Yes | No | Yes | Yes | Yes |
| GP wore a face mask | No | No | Yes | Yes | No |
| Risk contact score<sup>b</sup> | 2 | 2 | 2 | 2 | 2 |

<sup>a</sup>Combination of distance <1m and/or physical examination/blood drawn/injection; <sup>b</sup>Risk contact score (scale 0-3), one point for each: duration (1 point if $\geq 10$ minutes), distance (1 point for <1 meter and/or physical examination), GP with facemask (1 point for contact without a facemask, or contact $\geq 10$ minutes with a facemask)

No single type of contact was significantly associated with the risk of getting COVID-19 (**Table 2**); however, the trends indicated that contact with GP_X_, contact in the pre-symptomatic phase (data included for GP_X_ only, as GP_Y_ has not seen any patient in their pre-symptomatic phase), and contact ≥10 minutes increased the risk, while contact with a GP who was wearing a facemask decreased the risk (**Table 2**). All 22 identified COVID-19 cases had contact with a GP when the GP did not wear a mask, and/or when contact was ≥10 minutes. Importantly, contact <10 minutes with a GP wearing a facemask was significantly associated with a reduced risk (IRR 0.21; 95%CI 0.01-0.99) of COVID-19. When assessing the risk using a cumulative score (1 point for each: duration ≥10 minutes; distance <1 meter and/or physical examination; contact without a facemask or contact ≥10 minutes with facemask), trends indicated that increased exposure increased risk of COVID-19 (IRR per point 1.23; 95%CI 0.76-2.15).

**Table 2.** Incidence and incidence rate ratios (IRR) of COVID-19 cases (possible, probable and confirmed), by risk contact-event with SARS-CoV-2-positive general practitioners (GP).

| Type of risk contact | Exposure | Total<br>(n=89) | % | No case<br>(n=67) | % | Case<br>(n=22) | % | Incidence<br>rate ratios | 95%CI<br>lower | 95%CI<br>upper |
| --- | --- | --- | --- | --- | --- | --- | --- | --- | --- | --- |
| GP | GP Y | 46 | 51.7 | 36 | 53.7 | 10 | 45.5 | 1 (ref) | (ref) | (ref) |
|  | GP X | 43 | 48.3 | 31 | 46.3 | 12 | 54.5 | 1.28 | 0.55 | 3.04 |
| Phase of infection of GPs | Symptomatic | 20 | 22.5 | 16 | 23.9 | 4 | 18.2 | 1 (ref) | (ref) | (ref) |
|  | Pre-symptomatic | 69 | 77.5 | 51 | 76.1 | 18 | 81.8 | 1.30 | 0.49 | 4.51 |
| Duration | ≥10 minutes | 70 | 78.7 | 52 | 77.6 | 18 | 81.8 | 1 (ref) | (ref) | (ref) |
|  | <10 minutes | 18 | 20.2 | 15 | 22.4 | 3 | 13.6 | 0.65 | 0.15 | 1.91 |
| Facemask worn by GP | No facemask | 53 | 59.6 | 39 | 58.2 | 14 | 63.6 | 1 (ref) | (ref) | (ref) |
|  | Facemask | 31 | 34.8 | 24 | 35.8 | 7 | 31.8 | 0.85 | 0.32 | 2.05 |
|  | Unknown | 5 | 5.6 | 4 | 6.0 | 1 | 4.5 | - | - | - |
| Duration and facemask worn by GP combined<sup>a</sup> | Exposed (contact ≥10 minutes and/or GP not wearing facemask) | 75 | 84.3 | 53 | 79.1 | 22 | 100 | 1 (ref) | (ref) | (ref) |
|  | Unexposed (GP wore facemask and <10 minutes contact) | 14 | 15.7 | 14 | 20.9 | 0 | 0.0 | 0.21 | 0.01 | 0.99 |
| Risk contact score<sup>b</sup> | Per point (*continuous*) | NA | NA | NA | NA | NA | NA | 1.23 | 0.76 | 2.15 |

<sup>a</sup> IRR calculated using Haldane-Anscombe correction; <sup>b</sup> Risk contact score (scale 0-3), one point for each: duration (1 point if ≥10 minutes), distance (1 point for <1 meter and/or physical examination), GP with facemask (1 point for contact without a facemask, or contact ≥10 minutes with a facemask)

The results should be interpreted considering the following limitations. First, and due to the observational nature of our study, our case definitions are subject to potential misclassification. Our case definition for possible cases is very sensitive and includes symptoms that could also have been caused by pathogens other than SARS-CoV-2, e.g. seasonal influenza, which were still circulating at the time of the outbreak investigation. Conversely, the definition of confirmed cases relied on the availability of PCR test results. Although the majority of exposed patients (64/83) were tested, it is possible that asymptomatic cases or cases developing infection after the test were missed. Secondly, we cannot exclude that individual cases acquired their SARS-CoV-2 infection through a different exposure (i.e. not in the GP practice). However, as community transmission in the district was still very limited at the time (3, 4), we consider this possibility is unlikely. Third, we could trace 70%, but not all, of the listed GP contacts; this is a finding on its own. However, it does potentially introduced bias to the results, because we do not know if the contacts who were not reached had a specific profile irrespective of age or sex or whether the non-response was associated to being a case. Last, because of the limited cohort size, not all potentially confounding factors, such as patient characteristics (e.g. underlying illness) combined with possible differences in patient contact with GP_X_ or GP_Y_ could be considered in the statistical analysis.

### Conclusion

In conclusion, the use of facemasks for GPs and short consultation times helped limiting the spread of SARS-CoV-2 from GPs to their patients. Should a GP or other healthcare workers outside of a hospital setting with COVID-19 be shedding SARS-CoV-2 at the peak of infectiousness (i.e. shortly before or after symptom onset) then these types of infection prevention and control measures may prevent transmission to susceptible patients. Our investigation lends support to current guidelines from WHO and the ECDC, both recommending all healthcare workers to continuously wear a medical facemask at work especially in areas of community transmission (12-15).

## Data Availability

Detailed data are confidential and protected by German law. Anonymized data are available from the corresponding author upon reasonable request.

## Acknowledgments

We thank the Robert Koch Institute’s team supporting the interviews: Katharina Alpers, Nicu Anghel, Mona Askar, Eva Feuerhahn, Christina Frank, Jane Hecht, Alexandra Holzer, Anja Klingeberg, Teresa Nygren, Andreas Reich, Viktoria Schönfeld, Doreen Staat, Jan Walter, and especially Felix Moek for the organization. We further thank Merle Böhmer from the federal state public health authority and the local public health authorities, especially Clara Vicén Renner for carrying out several interviews. Last but not least, we very much appreciate and thank all participants of the study.

## Disclaimers

No conflict of interest reported. TSB and JB are fellows of the ECDC Fellowship Programme, supported financially by the European Centre for Disease Prevention and Control (ECDC). The views and opinions expressed herein do not state or reflect those of ECDC. ECDC is not responsible for the data and information collation and analysis and cannot be held liable for conclusions or opinions drawn.

